# Low circulating adropin levels in late-middle aged African Americans with poor cognitive performance

**DOI:** 10.1101/2021.12.09.21267550

**Authors:** Geetika Aggarwal, Theodore K. Malmstrom, John E. Morley, Douglas K. Miller, Andrew D. Nguyen, Andrew A. Butler

**Author notes:** Address for correspondence: Professor Andrew A. Butler, Department of Pharmacology and Physiology, Saint Louis University School of Medicine, St. Louis, MO, USA.

## Abstract

We recently reported accelerated cognitive decline in Europeans aged >70 years with low circulating adropin levels. Adropin is a small, secreted peptide that is highly expressed in the human nervous system. Expression profiling indicate relationships between adropin expression in the human brain and pathways that affect dementia risk. Moreover, increased adropin expression or treatment using synthetic adropin improves cognition in mouse models of aging. Here we report that low circulating adropin concentrations also associate with poor cognition (worst quintile for a composite score derived from the MMSE and semantic fluency test) in late-middle aged community-dwelling African Americans (OR=0.775, P<0.05; age range 45-65y, n=352). The binomial logistic regression controlled for sex, age, education, cardiometabolic disease risk indicators, and obesity. Previous studies using cultured cells from the brains of human donors suggest high expression in astrocytes. In snRNA-seq data from the middle temporal gyrus (MTG) of human donors, adropin expression is higher in astrocytes relative to other cell types. Advanced age suppresses adropin expression in all cell-types, with no effect of dementia status. In cultured human astrocytes, adropin expression also declines with donor age. Additional analysis indicated positive correlations between adropin amd transcriptomic signatures of energy metabolism and protein synthesis that are adversely affected by donor age. Adropin expression is also suppressed by pro-inflammatory factors. Collectively, these data indicate low circulating adropin levels are a potential early risk indicator of cognitive impairment. Suppression of astrocyte adropin expression in the brain is a plausible link between aging, neuroinflammation, and risk of cognitive decline.

## INTRODUCTION

Dementia is a blanket term used to describe the progressive, irreversible loss of intellectual abilities that result from neurodegenerative diseases^1-3^. Several types of dementia have been identified^3,4^. The most common form is Alzheimer’s disease (AD), which accounts for 60-80% of cases and is defined by the spread of extracellular amyloid-β plaques, intracellular tau tangles, and neuronal death^3,5^. Risk for dementia increases with advanced age; about 1-in-3 people aged 80 years of older develop dementia^3^. Mild-cognitive impairment (MCI) is often the first sign of dementia, with one-third of people with MCI developing AD within 5 years^3^. In the absence of widely available effective therapies that prevent or delay the progression of dementia, the prevalence of this dementia is predicted to increase as gains in life expectancy result in aging of the population.

Recent advances in neuroimaging and blood-based biomarkers have improved dementia diagnosis and monitoring of disease progression^5-8^. Some of the blood-based biomarkers reflect the accumulation of amyloid and neurofibrillary tangles in the brain^5,6,8^. Increased circulating levels of another biomarker (neurofilament light chain, or NfL) reflect axonal damage due to traumatic brain injury, inflammatory processes, or cerebrovascular diseases^7^. Recent trials of antibody therapies that target beta amyloid indicate that delaying cognitive decline in patients with AD may be possible, although the risk of adverse side effects (brain swelling, hemorrhaging) is significant^9-12^. These results nevertheless suggest therapies targeting specific disease-driving pathologies identified with clinical testing could be achievable. However, further research is needed to reduce risk associated with these treatments and discover new modifiable risk factors for dementia.

The secreted peptide adropin could be a novel risk indicator for accelerated cognitive decline. Low circulating levels of adropin associated with accelerated cognitive decline in community-dwelling old individuals in the Multidomain Alzheimer’s Preventive Trial (MAPT)^13^. The pathologies that underlie the relationship between circulating adropin levels and cognition are not known. However, expression profiling of the mRNA encoding adropin in humans and nonhuman primates and measurements of adropin protein expression in mice suggest it is abundant in the nervous system^14-16^. In all three species, adropin expression clusters with gene networks implicated in neurodegenerative conditions^14,15^. For example, in human postmortem brain samples strong positive correlations are observed between adropin expression and gene networks involved in mitochondrial function, synaptic plasticity, and cerebrovascular function^14^. These processes are relevant to maintaining brain health during aging^17^.

Studies using rodent models support the hypothesis that high adropin activity preserves cognitive ability in aging. Adropin expression in the mouse and rat brain declines with aging^14,18^, while adropin treatments improve cognition in 18-month-old male mice^14,19^. This outcome potentially involves direct and indirect actions on neurons. Adropin is neurotrophic when applied directly to cultured neurons^14^, while also preserving cerebrovascular function in situations of ischemia^19-22^. Adropin could thus support synaptic plasticity in the brain while also preventing neurovascular diseases thought to contribute to cognitive decline^23^. However, whether the results from mouse studies are translatable to the human condition has not been determined.

To date, there has been only one study investigating the relationship between circulating adropin concentrations and cognitive impairment in humans^13^. The primary objective of the current study was therefore to determine whether the relationship between circulating adropin and cognition replicates in different populations. A secondary objective was to further investigate the expression profile of the Energy Homeostasis Associated (ENHO) mRNA transcript that encodes adropin in the human nervous system.

## RESULTS

### Low serum adropin concentrations in late middle-aged African Americans with poor cognition

Mean age, sex, BMI, and serum adropin concentrations of the subjects used for this study are shown in **supplementary table 1**. We first tested the hypothesis that low circulating adropin concentrations associate with poor cognition. Poor cognition was defined as scoring in the worst quintile for a composite of two tests (MMSE, ANT). Test scores, age, years of education (self-reported), and serum adropin concentrations in the poor cognition group relative to the 2^nd^-5^th^ quintile group are shown in **Table 1**. People defined as having poor cognition had on average, fewer years of education and were slightly (2 years) older. They also had significantly lower serum adropin concentrations. The difference remained significant after controlling for the potential effects of age, obesity, glucose control, dyslipidemia, and sex on serum adropin concentrations (**Table 2**).

**Table 1.** Physical characteristics and plasma variables (unadjusted) for study participants with poor cognition. Poor cognitive performance was defined as scoring in the worst quintile for a composite of two tests (MMSE, ANT). Data are reported as mean ± std. deviation, with sample sizes shown in brackets. The sample size for the sum of the low quintile and 2^nd^-5^th^ quintiles may not match owing to missing data. The *p* values shown are from comparisons using Mann-Whitney U test. The Chi-Square test was used to assess the distribution of sex between the two groups. Not statistically significant – n.s. Serum leptin concentrations are estimated marginal means adjusted for sex.

| Variable | Worst quintile (n=67) | 2<sup>nd</sup>-5<sup>th</sup> quintiles (n=262) | *p* value |
| --- | --- | --- | --- |
| % Female | 62.7% | 67.9% | n.s. |
| MMSE+ANT (Z) | -2.41 ± 1.16 | +0.63 ± 1.07 | <0.001 |
| MMSE | 24 ± 3 | 29 ± 1 | <0.001 |
| ANT | 14 ± 4 | 21 ± 6 | <0.001 |
| Age (years) | 58 ± 4 | 56 ± 4 | 0.005 |
| BMI (kg/m<sup>2</sup>) | 30.5 ± 7.0 | 31.6 ± 7.0 | n.s. |
| Years of education | 10 ± 3 | 13 ± 3 | <0.001 |
| Fructosamine (μmol/L) | 277 ± 76 | 263 ± 62 | n.s. |
| TG (mg/dL) | 146 ± 80 | 142 ± 71 | n.s. |
| Adiponectin (μg/ml) | 9.1 ± 6.6 | 8.0 ± 5.0 | n.s. |
| Leptin (ng/ml) | 42.8 ± 3.9 | 47.5 ± 2.1 | n.s. |
| Adropin (ng/ml) | 2.87 ± 1.22 | 3.34 ± 1.48 | 0.036 |

**Table 2.** Serum adropin concentrations adjusted for age, BMI, leptin, adiponectin, fructosamine, and TG in male and female study participants scoring in the low quintile for cognition. Data shown are estimated means ± SE, significant differences between low_quintile and 2^nd^-5^th^_quintiles are indicated by asterisks (*p<0.05; **, p<0.01).

| **Sex (N)** | **Worst Quintile (N)** | **2<sup>nd</sup> - 5<sup>th</sup> Quintile (N)** |
| --- | --- | --- |
| Male<br>(110) | 2.58 ± 0.30<br>(25) | 3.35 ± 0.18*<br>(85) |
| Female<br>(218) | 2.89 ± 0.22<br>(43) | 3.40 ± 0.12*<br>(175) |
| All<br>(328) | 2.73 ± 0.18<br>(68) | 3.37 ± 0.10**<br>(260) |
| Covariates with p<0.05: Leptin (p<0.05); Adiponectin (p<0.005). |  |  |

A decline in the prevalence of poor cognition as adropin values increase was also observed using binomial logistic regression (odds ratio [OR]=0.772, p=0.014, 95% CI 0.629-0.948, n=338). Several studies have observed relationships between circulating adropin and indices of obesity and dysregulated glucose and/or lipid metabolism^24,25^. A follow-up analysis included additional covariates that might affect circulating adropin levels or cognition (age, sex, years of education, BMI, adiponectin, leptin, fructosamine, and TG). Of note, weak correlations were observed between serum levels of adropin and adiponectin (*ρ* = 0.198, p<0.001), fructosamine (*ρ* = 0.128, p<0.05), and TG (*ρ* = -0.121, p<0.05). However, including these covariates did not affect the relationship between adropin and poor cognition (OR=0.775, p<0.05) (**Table 3**). Of the other variables included in the analysis, only years of education was a significant predictor of poor cognition (OR=0.635, p<0.001) (**Table 3**).

**Table 3.** Risk of poor cognitive performance in relation to serum adropin concentrations in the AAH cohort.

| **Variable** | **Odds Ratio (95% CI)** | ***p* value** |
| --- | --- | --- |
| **Age (years)** | 1.060 (0.987-1.139) | 0.108 |
| **Sex (category)** | 1.026 (0.428-2.460) | 0.906 |
| **Education (years)** | 0.635 (0.543-0.742) | <0.001 |
| **BMI (kg/m<sup>2</sup>)** | 0.993 (0.931-1.059) | 0.826 |
| **Adiponectin (µg/ml)** | 1.035 (0.975-1.099) | 0.258 |
| **Leptin (ng/ml)** | 0.995 (0.981-1.009) | 0.446 |
| **Fructosamine (µmol/L)** | 1.002 (0.997-1.006) | 0.420 |
| **TG (mg/dL)** | 1.002 (0.997-1.006) | 0.461 |
| **Adropin (ng/ml)** | 0.775 (0.611-0.982) | 0.035 |

Serum adropin concentrations did not correlate with circulating markers of inflammation (data not shown). High neurofilament light chain (NfL) levels can indicate increased risk of dementia ^26^. However, there was no correlation between serum adropin and NfL concentrations (data not shown).

### Adropin expression in human astrocytes correlates with donor age and metabolic activity

We have reported high adropin expression in cultured human astrocytes relative to other cell types isolated from brain tissue samples^14^. We used snRNA-seq data to compare relative expression levels and fraction of cells within each population in the human MTG in a reference neurotypical population of males and females (aged 18-68 years, n=5) (**Fig 1A, B**) and an older cohort of males and female with mixed dementia status (aged from 65 to 90+ years, n=84) (**Fig. 1C, D**) ^27^. The data from this study indicates astrocytes are a major site of ENHO expression relative to other cell types, irrespective of age (**Fig. 1A-E**). Oligodendrocyte precursor cells (OPC) also exhibit comparable levels of ENHO expression, but in a smaller fraction of this population (**Fig. 1E**). A few populations of neurons appear to exhibit a compatible fraction of cells showing ENHO expression (e.g., Pax6^+ve^ and Vip^+ve^ neurons); however, the relative level of expression is lower compared to astrocytes. There appears to be a reduced expression in the older cohort, irrespective of age, sex, and dementia status (**Fig. 1E, supplementary figures 1 and 2**). Moreover, the distinction between the two non-neuronal populations with high expression (astrocytes>neurons) may increase with age.

**Figure 1.**
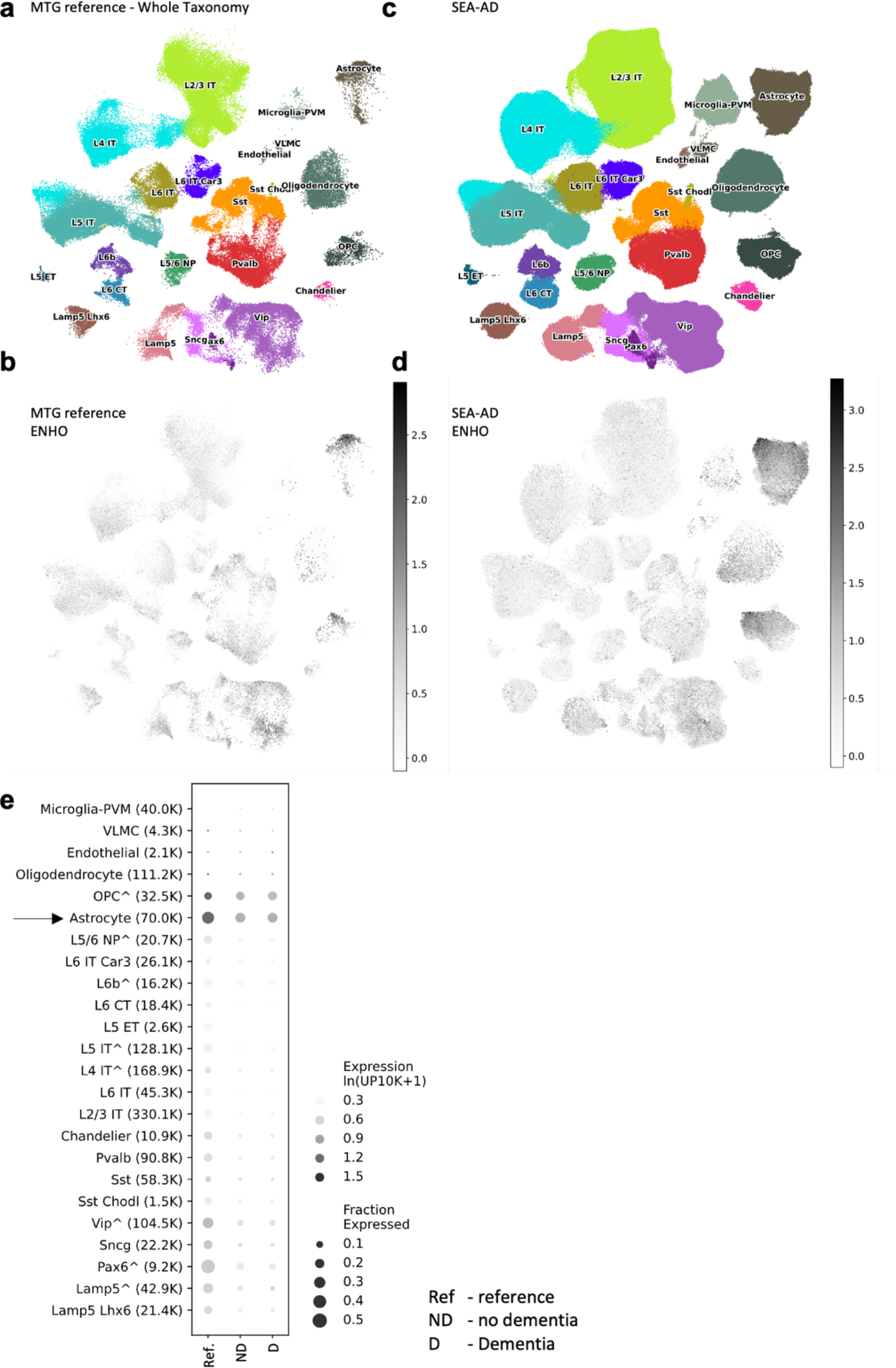
Comparison of ENHO expression in nuclei isolated from MTG of a neurotypical reference population and aged donors with or without dementia. UMAP representations of cell population using color-coding to identify cell subclass/supertype (A, C) or relative ENHO gene expression (B, C). The data shown are for the MTG reference group (A, B; n=5) or donors with a range of AD neuropathologic change (ADNC) (C, D, n=84). (E) Dotplot comparison of ENHO gene expression between the MTG reference group (n=5) and donors with no dementia (n=42) or dementia (n=42). These data were extracted from the Seattle Alzheimer’s Disease Brain Cell Atlas.

In astrocytes isolated from human donors, ENHO expression levels also appear to decline with donor age (**Fig. 2A**). While astrocytes from male donors exhibit a greater range of expression, donor sex does not significantly affect expression levels (**Fig. 2B**). ENHO expression adjusted for donor age was not significantly different (estimated marginal means and std. error for males, 3.01±0.351; for females, 2.11±0.35; p=0.108).

**Figure 2.**
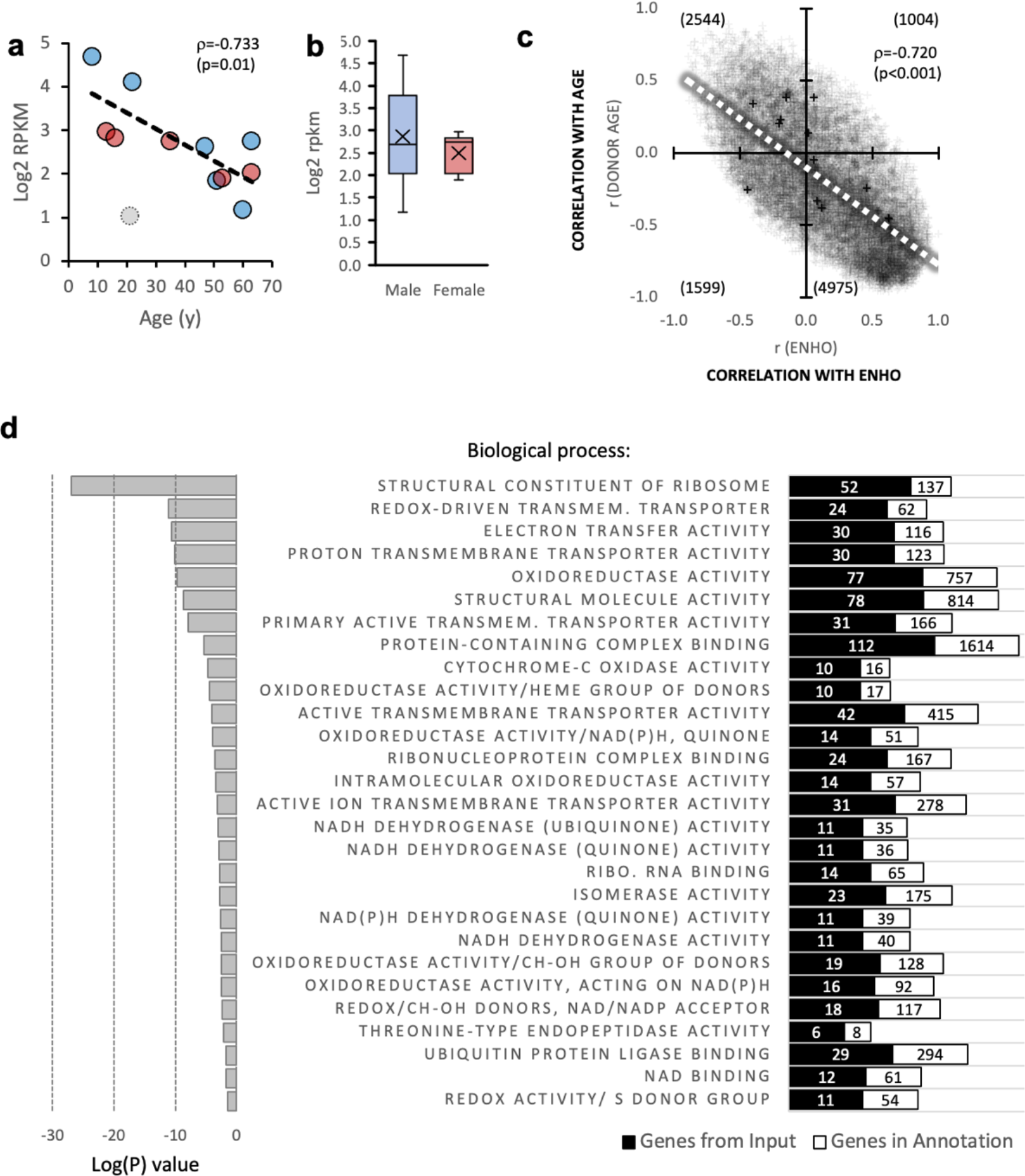
Expression of the adropin (ENHO) transcript in isolated astrocytes. (A) ENHO expression as a function of donor age. The correlation coefficient shown is after removal of one outlier (open circle); including the outlier results in a trend for an association (*ρ*=-0.529, p=0.077). Data from male donors is shown in blue, red for females. (B) ENHO expression levels in astrocytes from male or female donors (n=6/group). (C) Relationship between the pair-correlation matrices for transcripts paired with donor age (y-axis) or ENHO (x-axis). Most transcripts (50%) are in the quadrant that correlate positively with ENHO and negatively with age. (D) Results from the gene enrichment analysis showing cellular pathways correlating positively with expression of ENHO (adropin).

The results from snRNA-seq and cultured astrocytes nevertheless suggest that ENHO expression declines with aging. A similar aging effect has been reported at the level of adropin protein expression in cortical lysates from shorter-lived rodent models^14,18^.

To identify gene networks co-expressed with ENHO in cultured astrocytes and how this relationship might be affected by donor age, a paired regression approach was used^15^. Correlation coefficients were calculated by comparing all transcripts against ENHO or donor age. Comparison of the correlations between all transcripts and either ENHO or donor age suggested an interaction (**Fig. 2C**). Transcripts that correlate positively with ENHO correlated inversely with donor age. ENHO transcript levels thus appear to be co-regulated with clusters of genes affected by donor age.

We next determined whether transcripts co-regulated with ENHO are enriched for specific pathways. To control for donor age, a paired correlation matrix using the ENHO transcript as bait was re-calculated using donor age as a covariate. This approach identified 752 genes with a paired correlation coefficient with ENHO of >0.7 (supplemental data file, **supplementary table 2**). Gene ontology analysis indicated highly significant enrichment for pathways related to mitochondrial oxidoreductase reactions and energy-demanding synthesis of micro- and macromolecules (**Fig. 2D**, supplemental data file, **supplementary table 3**). Repeating the analysis using a simple paired correlation without donor age as a covariate yielded a similar outcome (supplemental data file, **supplementary table 3**). While this approach identified more genes (1627 genes), transcripts encoding proteins involved in similar metabolic processes were enriched, albeit with reduced statistical significance.

We next compared transcripts with an R>-0.7 when paired with either ENHO transcript level (2276 transcripts) or an R<-0.7 paired with donor age (1627 genes). There was significant overlap between the population (826 genes) (supplemental data file, **supplementary table 4**). The 826 genes common to both groups were also enriched for pathways involved in processes related to macromolecule production and turnover (intracellular transport, protein localization, protein synthesis, autophagy, lipid synthesis, glucose metabolism). The decline in ENHO expression with donor age may thus reflect a general reduction of metabolic processes and cellular energy use in astrocytes.

For transcripts correlating negatively with ENHO, fewer met the selection condition (420 in age-adjusted matrix, 248 in the simple matrix (supplemental data file, **supplementary table 5**). Significant enrichment was only observed for the age-adjusted gene set, and primarily identified pathways active in the cell nucleus. Comparison of the genes identified by simple correlations as correlating negative with ENHO and positively with age was not informative with respective to pathway enrichment.

### Regulation of astrocyte adropin expression by inflammation

Adropin expression in the rodent brain is suppressed with aging, and correlates inversely with inflammatory markers^14,18^. Experiments using astrocytes isolated from either human or mouse donors (GSE147870) suggest ENHO expression is suppressed by the TLR3 agonist Poly I:C, and by TNFα (**Fig. 3A-F**). A two-way ANCOVA (independent variables: treatment, donor species) indicated an inhibitory effect of Poly I:C and TNFα irrespective of the donor species (treatment effect, p<0.05). Adropin is however not regulated by hypoxia in astrocytes isolated from mice or human brain samples (**Fig. 3**).

**Figure 3.**
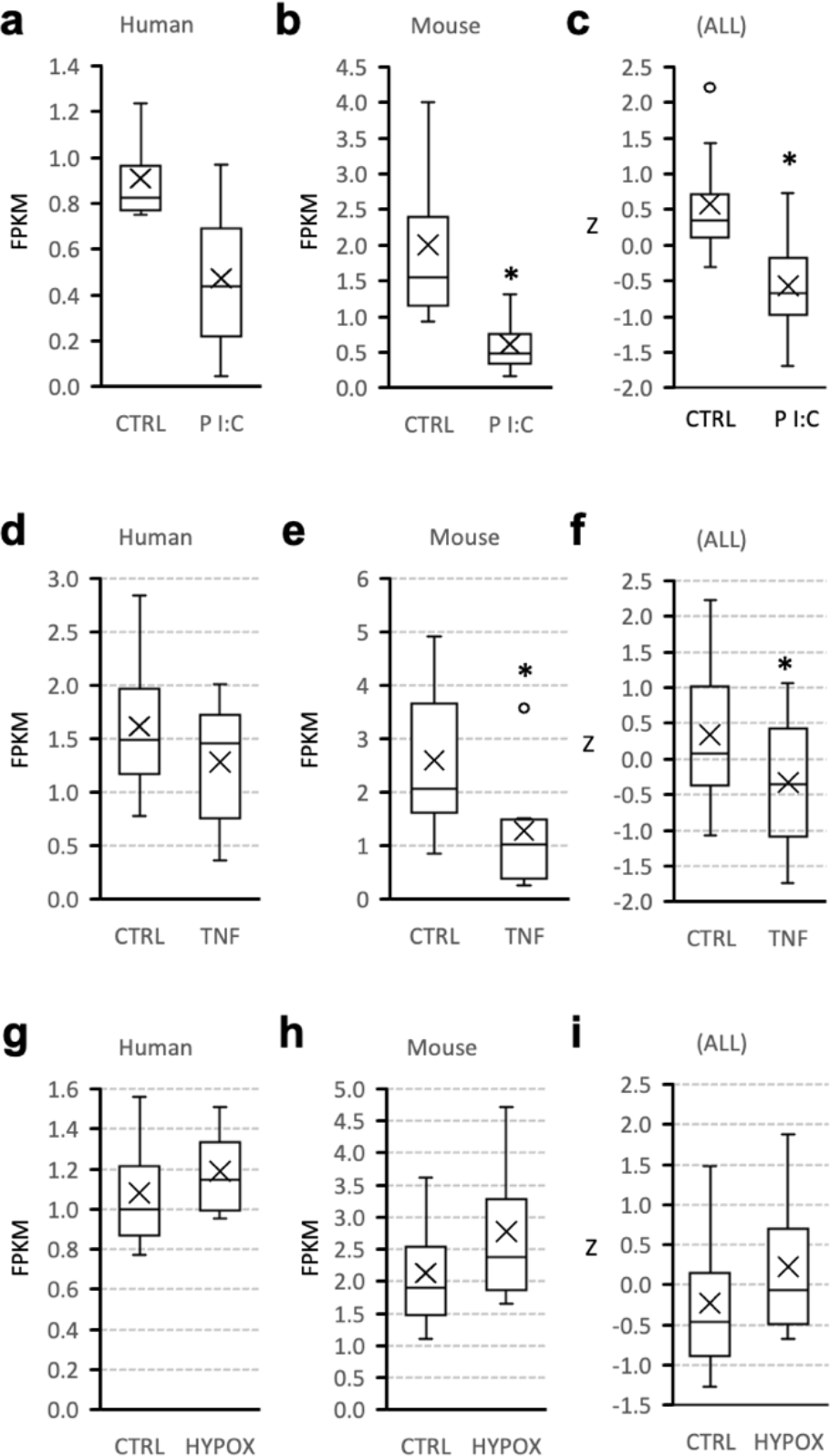
Expression of the adropin (ENHO) transcript is suppressed by inflammatory signals. Regulation of ENHO expression in astrocyte cultures from human or mouse donors by the TLR agonist Poly I:C (A-C, 200 μg/ml for 72h; n=4 per treatment group within species), TNFα (D-F, 30 ng/ml for 48h; n=9 per treatment group for human donors, n=6 per treatment group for mouse donors), or hypoxia (G-I, 1% O_2_ for 72h; n=4 per treatment group within species). * p<0.05 vs. control (CTRL) by Mann Whitney U-test (a, b, d, e, g, h) or 2-way ANOVA (c, f, i). The box and whisker charts show the distribution of data by quartiles, and highlight the mean (X) and outliers (circle).

## DISCUSSION

Our analysis of circulating adropin concentrations in the MAPT study indicate low circulating adropin concentrations associate with increased risk of accelerated cognitive decline in people aged over 70 years ^13^. However, human transcriptome data indicated that adropin could provide an indication of cognitive performance earlier in life ^14^. Indeed, expression of the adropin transcript in the brain appears to peak in the first decade of life^14^. This investigation therefore began by testing the hypothesis that low serum adropin concentrations would indicate poor cognitive performance using a younger cohort.

The first part of study used a well-characterized, population-based cohort of community dwelling African American individuals aged between 45-65 years. Study participants with poor cognition, defined in this study as the worst quintile for a composite test, had significantly lower circulating adropin concentrations. Binomial logistic regression further indicated that increasing circulating adropin concentrations associate with reduced prevalence of poor cognition. These findings extend upon our recent findings indicating that adropin signaling functions, either directly or indirectly, to support cognitive function. Furthermore, they suggest that an association between circulating adropin concentrations and poor cognitive performance may be evident earlier in the life cycle.

The rationale for this study was based on our previous reports using human expression profiling data and mouse models of aging^14^. Expression of the transcript encoding adropin in post-mortem brain samples correlates positively with pathways that affect risk of cognitive decline (mitochondrial function, synaptic plasticity, vascular function, inflammation)^14,18,21,22^. Moreover, experiments using C57BL/6J mice suggest high adropin expression prevents or delays cognitive decline with natural aging ^14^ or obesity-related metabolic dysregulation (hypercholesterolemia, type 2 diabetes)^28^. This could involve a direct trophic action to support neuronal networks^14^. However, adropin could also act on the brain vasculature to preserve blood-brain barrier integrity and reduce risk of ischemia ^19,21,22^. Further studies are needed to determine the pathologies underlying the association between circulating adropin levels and cognition.

The results from this study are a significant extension of our analysis of circulating adropin in the MAPT study^13^. In that study, circulating adropin concentrations did not correlate with cognitive ability at the time of sampling, but correlated with subsequent cognitive decline. The reasons for the disparity are not clear, but it is important to note the differences between the two studies. The MAPT study involved older Europeans who had access to universal healthcare. In contrast, the AAH involved self-reported African Americans living in the inner city or suburban St. Louis who had likely experienced racial disparities in health care^29^. The MAPT used a comprehensive analysis of cognitive function, while the current study was limited to the MMSE and ANT. Indeed, a weakness of the current study is that it only used two cognitive tests. The current study was also not designed to investigate cognitive decline. While suggesting an association between serum adropin levels and cognition, the long-term clinical significance with respect to risk of further cognitive decline in younger cohorts are not known and require further study.

Using the exploratory resources from the Seattle Alzheimer’s Disease Brain Cell Atlas consortium, we investigated the distribution of ENHO expression in specific cell population in the MTG. We had previously observed relatively high ENHO expression in mature astrocytes compared to other cell populations (neurons, oligodendrocytes, glial cells, endothelial cells) maintained in culture media after isolation from human brain samples ^14^. The current results from the analysis of snRNA-seq data indicate a similar distribution in the brain. These results further suggest that astrocytes are a major site of ENHO expression in the human brain. A caveat is that the current study was limited to one region of the brain (the MTG). It is possible that the distribution between cell types differs between brain regions. Further examination of the distribution of adropin between cell-types within different brain regions will be needed to support the current findings.

The source(s) of circulating adropin remains to be identified. The brain and (to lesser degree) liver have been identified as major sites of expression ^14-16,30,31^. In a diurnal NHP (the baboon), brain adropin expression expressed as fragments per kilobase of transcript per million reads mapped, or FPKM) is orders of magnitude higher relative to non-neural tissues and exhibits a circadian profile with peaks during the daytime ^15^. Circulating adropin concentrations in another NHP model, the rhesus macaque, also appear to exhibit a diurnal profile with higher levels in the daytime ^15^. Collectively, these results suggest that adropin could be a circadian-controlled neuropeptide, with levels in the circulation reflecting expression in the nervous system. However, further study of the source of circulating adropin are needed. It is however important to note that comparison of RNA-seq data between cell and tissue types should be interpreted with caution. Expression levels reported at FPKM or TPM (transcripts per million) don’t account for differences in RNA content between cell- and tissue-types^32^. Moreover, for human tissue samples the variability in interval between death and tissue preservation can affect both RNA quality and yield^33^.

The initial studies investigating circulating levels of adropin resulted from data suggesting adropin^1-76^ is a prohormone with a signal peptide (adropin^1-33^) and a secreted peptide (adropin^34-76^) ^30^. Indeed, adropin is often described as a liver-secreted ‘hepatokine’ ^25,34^. However, a later study refuted the finding that adropin is secreted, describing instead a membrane-bound protein primarily expressed in the brain ^16^. This conclusion is supported by data from AlphaFold’s transmembrane protein structure database, which predicts a transmembrane region near the N-terminus. We have observed adropin secretion by human iPSCs can be detected using the enzyme immunoassay used for this study in conditioned media (Aggarwal and Nguyen, unpublished data). This result is predicted based on the results from experiments that used an C-terminal epitope-tagged adropin fusion protein which indicated a portion of the prohormone is secreted into the media by cultured cells, and into the mouse circulatory system when expressed in the liver ^30^. These results suggest an as-yet unidentified mechanism for releasing a portion of the adropin prohormone from the cell.

A strength of the brain cell atlas dataset is the involvement of older subjects with a range of AD pathology. While ENHO expression appears to decline with aging, expression does not appear to be markedly different between people with or without dementia. Indeed, comparisons between the 84 older subjects and the reference group suggests that age may have an inhibitory effect on expression.

In shorter-lived mice and rats, adropin protein levels in the brain decline between 3 and 18 months of age^14,18^. Preventing this decline, or treatment using synthetic adropin peptide, improves spatial learning and memory^14^. However, whether the decline is sufficient *per se* to cause cognitive impairment is not known. While studies have examined how adropin therapy affects cognition in mouse models, there have been no studies to determine whether deficiencies in adropin expression accelerate cognitive decline in aging.

ENHO expression in cultured human astrocytes also correlates inversely with donor age, which is consistent with the results from the analysis of expression in the MTG using snRNA-seq. This relationship appears to correlate with changes in metabolic activity. Specifically, ENHO expression adjusted for donor age correlates positively with cellular pathways involved in energy-demanding cytosolic processes involved in macromolecule synthesis and turnover. ENHO expression also correlates with the mitochondrial process needed to provide the energy for these processes. High adropin expression in young astrocytes thus appears to correlate with higher metabolic activity and energy demand, with energy production presumably required for the synthesis of macromolecules (RNA, proteins, lipids).

This result is important, as it is consistent with the relationships observed between ENHO expression and gene clusters in RNA-seq data derived from human post-mortem brain samples^14^. High adropin expression in the brain may therefore reflect relatively high levels of metabolic activity. Experiments using mice suggest that this relationship could be causal, as adropin treatment enhances the expression of similar pathways in the mouse brain^14^. Further studies are clearly needed to determine whether circulating adropin levels correlate with brain ENHO expression and with pathways involved in energy utilization.

In rodent models of aging, a decline in adropin protein levels is observed in brain lysates^14,18^. Neuroinflammation is one of the key mechanisms identified in neurodegenerative diseases^35,36^. Here we observed evidence of a direct response of ENHO transcript expression in astrocytes to TNFα and a TLR3 agonist. While the effect is observed in astrocytes from human and mouse donors, the effect appears more robust in the latter. It is important to note that the astrocytes used for this study were isolated from developing brains^37^. The relevance to the conditions observed in the aginh brain is therefore unclear. However, the results are significant in suggesting a direct inhibitory effect of inflammatory signals on adropin expression in astrocytes.

It important to note that ENHO expression was not lower in older people diagnosed with dementia compared to those who did not exhibit dementia. The expression profile of ENHO in older people categorized by BRAAK staging also indicated no decline with increasing severity. This suggests that low ENHO expression *per se* is unlikely to be a primary driver of dementia in older individuals. Rather, low adropin expression levels in the brain, and levels of circulating adropin, could facilitate disease progression. In this model, reduced neuroprotection due to low expression would have less constraint on disease progression driven by other genetic and environmental risk factors. This model is consistent with data from experiments using mice that over express adropin ^14,19,21,28^. Adropin-deficient mice are more susceptible to cerebrovascular ischemia ^19,21^. However, experiments using mouse models are needed to explore the relationship between low adropin expression and cognitive decline driven by other risk factors for dementia. Furthermore, the relationship if any between adropin expression in the brain of young to middle-aged people with cognitive functions is not known, and was not addressed in this study.

There are limitations to the current study. These include the use of a cohort housed in a single geographic location. The study limited to a single racial group (African Americans), albeit from two socioeconomically diverse geographic areas. Another limitation was that the inclusion criteria for the AAH study included a MMSE score of ≥16 ^29,38^. The study population therefore does not include late middle-aged people with dementia. The ceiling effect observed with the MMSE is also a limitation; it has a maximum score of 30 which was achieved by nearly a third of the subjects included in the study. And as already stated, only two tests were used to assess cognitive function, and there was some inconsistency in the correlations between the two tests. These results should therefore be considered preliminary. Additional studies are clearly needed to explore the relationships between circulating adropin concentrations and risk of dementia.

In summary, the results of the current study indicate that low circulating adropin concentrations in late middle-aged people associate with poor cognitive function. Further studies are needed to determine whether circulating adropin concentrations correlate with risk for cognitive decline with aging and impaired glucose homeostasis.

## METHODS

### African American Health (AAH)

AAH was a population-based, community-dwelling cohort of 998 self-identified African Americans in 2000–2001 in the Saint Louis metropolitan area^29,38-40^. Recruitment used a multi-stage probability sampling methodology aimed at selecting approximately equal numbers of participants from inner-city and near suburban neighborhoods northwest of Saint Louis City. The study procedures were reviewed and approved by the Institutional Review Board at Saint Louis University and were performed in accordance with the ethical standards laid down in the 1964 Declaration of Helsinki and later amendments. All participants provided written consent for study, including blood draw.

Inclusion criteria for AAH included self-reported Black or African American race; birth dates between 01/01/1936 and 12/31/1950; standardized Mini-Mental Status Examination (MMSE) scores ≥ 16; and willingness to sign an informed consent form^29,38-40^. Participants initially completed in-home interviews and assessments at baseline, and at years 3 and 9 the recruitment proportion (participants/enumerated eligible persons) was 76%. Self-reporting bias was minimized using validated and reliable criteria for collecting self-reported data. A subset of participants agreed to donate blood; serum samples were stored at −80 °C for laboratory analyses. This was a community-based study; fasting was not required at the time of blood collection to encourage participation. Age and years of education are self-reported. For age, participants were asked to report their age and date of birth; mismatches between the two answers resulted in a query for clarification.

The current study established baseline fed serum adropin concentrations for 352 of the participants for which serum was still available. These values were compared to baseline cognitive data and serum markers of metabolic homeostasis and immune function.

### Mental status

Cognitive performance was assessed by the mini-mental state examination (MMSE) and one-minute animal naming test of verbal fluency (ANT) performed at the same time during the at-home evaluations. The MMSE is a screening tool for detecting cognitive deficits in dementia and delirium^41^. The MMSE is comprised of 11-questions that test five areas of cognitive function (orientation, registration, attention and calculation, recall, and language), with a maximum score of 30. There is a ceiling effect with the MMSE test which does not occur in the ANT test, which is a limitation of this approach. Out of 352 subjects, 142 had an MMSE score of 30.

A composite cognitive test score was calculated using the sum of the Z scores for the results of both tests. MMSE scores of ≤23 or ANT scores of ≤15 predict risk for dementia ^42,43^. The ANT assesses memory recall and verbal fluency ^43^.

For this study, poor cognitive performance was defined as scores in the worst quintile (“worst_quint”) for the composite score (MMSE+ANT_worst_quint) (**supplementary figure 3A**). For the composite score, both MMSE and ANT scores were significantly lower in the MMSE+ANT_worst_quint group relative to the 2^nd^ to 5^th^ quintile group (**supplementary figure 3A**). MMSE and ANT scores exhibited a modest correlation (*ρ* =0.333, p<0.001, n=338) (**supplementary figure 3B**). However, the composite score highly correlated with the results from both tests (MMSE, *ρ* =0.752, p<0.001; ANT, *ρ* =0.832, p<0.001). (**supplementary figure 3C, D)**.

### Laboratory procedures

Laboratory analyses used blood collected from a subset of AAH participants within approximately 2 weeks of the at-home assessment of mental status. Serum adropin concentrations were measured using an enzyme immunoassay kit from Phoenix Pharmaceuticals, Inc. (cat. no. EK-032-35) following the manufacturer’s protocol. Assay sensitivity reported by the manufacturer is 0.3 ng/ml, with a linear range from 0.3 to 8.2 ng/ml. Pilot experiments determined a 1:5 dilution of serum produced values within the linear range. Values <0.3 ng/ml were described as ‘low’ (n=1) and arbitrarily assigned a value of 0.15 ng/ml (half of the lowest detectable value). Values >8.2 ng/ml were described as “high” and arbitrarily assigned a value of 8.5 ng/ml (n=2).

Adropin assays were performed in triplicate using fifteen 96-well plates. The serum used for this study had been through several freeze/thaw cycles for previous analysis. Plate controls included an ‘in-house’ human serum sample, and controls provided by the assay manufacturer. The %CV for the plate controls were 11% and 14%, respectively. The initial criterion for inclusion was based on the coefficient of variation (%CV, SD divided by the mean of three replicates). For samples with %CV>20, the ‘outlier’ was removed. Data from triplicates with no clear outlier (5 samples) were excluded. The mean, median, and standard deviation (SD) for the 347 samples used in the analysis were 3.17, 3.11, and 1.35 ng/ml, respectively.

The procedures used to measure serum indices of metabolic control and inflammation have been described previously ^44,45^. Briefly, fructosamine was measured as previously described^46^. The fructosamine test measures average glucose levels over two to three weeks prior to testing, and can be used in situations where the hemoglobin A1C test is not reliable (hemolytic or sickle-cell anemia, renal disease, liver disease, HIV infection, recent blood loss) ^47^. Fructosamine values have also been reported to more consistently correlate with continuously monitored glucose values than A1C^47^. As detailed medical evaluations were not available, fructosamine values were used to estimate of averaged glucose levels over multiple days. Fructosamine values in people without diabetes range from 175 to 285 μmol/L; values of 286 μmol/L or higher indicate impaired glucose homeostasis and elevated blood glucose levels.

The soluble Interleukin-6 Receptor (sIL-6R) predominantly results from cleavage from cell-surface IL-6R, and is involved in IL-6 trans-signaling, a process which results in most cell-types becoming responsive to IL-6 ^48^. IL-6 circulates bound to sIL-6R, and the levels of sIL-6R are increased in inflammatory states ^48^. The levels of sIL-6R were determined using ELISA (ICN-Biomedicals, Costa Mesa, CA; intra-assay CV 5.0%; interassay CV 5.9%). High levels of other soluble cytokine receptors and C-reactive protein (CRP) were previously shown to associate with poor physical performance in the AAH ^49^. ELISA kits (BioSource, Camarillo, CA) were used for measuring soluble Tumor Necrosis Factor Receptor-1 (sTNFR1) (intra-assay CV 4.1%, interassay CV 7.3%) and soluble Tumor Necrosis Factor Receptor-2 (sTNFR2) (intra-assay CV 5.1%, interassay CV 8.6%). C-Reactive Protein (CRP) was measured using a High-Sensitivity Enzyme Immunoassay (hsCRP ELISA) (MP Biomedicals, Orangeburg, NY; intra-assay CV 4.5%, interassay CV 4.1%).

### Expression profiling

ENHO expression in human brain samples was assessed using data from the Seattle Alzheimer’s Disease Cell Atlas consortium (http://portal.brain-map.org/explore/seattle-alzheimers-disease)^27^. ENHO expression was compared in single nuclei isolated from the middle temporal gyrus (MTG) of 84 older individuals (33 males, 51 females; mean and SD of age at time of death, 88 years, 8years; range 65 to 90+ years) and a younger neurotypical reference group (male and females aged 18-68 years, n=5).

To investigate the regulation of ENHO expression in cultured astrocytes, RNA sequencing (RNA-seq) data from two published experiments was downloaded from the Gene Expression Omnibus (GEO) ^50^. Zhang *et al*. (GSE73721) used an immunopanning method to isolate astrocytes from juvenile (8-18y of age) and adult (21-63y of age) human brain tissue samples extracted during neurological surgery ^51^. The samples were from the healthy temporal lobe cortices and were rapidly processed following surgical removal. For the ‘immunopanning’ protocol, single-cell suspensions of dissociated tissues were incubated sequentially in a series of antibody-coated Petri dishes, with an anti-HepaCAM antibody used to bind and extract astrocytes.

The RNAseq data from the study of Zhang *et al*.^51^ were used to identify gene networks correlating with *ENHO* expression in human astrocytes. These were identified using a paired partial correlation analysis as previously described ^14,15^; donor age was used as a covariate in calculating the correlation matrix. Only genes with a mean FPKM of >1 were included in the analysis. Correlation coefficients of >0.7 or <-0.7 were used as selection criteria. The ToppGene Suite was used to identify gene networks overrepresented in genes identified as correlating positively or negatively with the ENHO transcript encoding adropin. Li et al. (GSE147870) applied a similar protocol to extract astrocytes from human and mouse brains^37^.

### Statistical analysis

Data were analyzed using IBM SPSS Statistics version 19.0 (IBM Corp., Somers, NY). Unadjusted data are presented as mean ± standard deviation. Comparisons of unadjusted data between two groups (e.g., worst quintile vs. 2^nd^-5^th^ quintiles) used a nonparametric test (Mann-Whitney U test, two-sided). When needed, data from ANCOVA adjusted for covariates (indicated in the text) are reported as estimated marginal means ± standard error. Binomial logistic regression was used to estimate the relationship serum adropin concentrations and risk of poor cognitive performance. Poor cognitive performance was defined as scoring in the low quintile for the composite score (MMSE+ANT_worst_quint). Modelling of the relationships between serum adropin concentrations and indices of glycemic control (fructosamine, leptin, adiponectin), lipid metabolism, and inflammation used Spearman’s correlation coefficient (*ρ*).

## Data availability

Access to AAH study data is available upon request. The RNA-seq data sets from cultured astrocytes are available from the NCBI Gene Expression Omnibus (GEO) website.

## Code availability

No code was generated for this study.

## ACKNOWLEDGEMENTS

AAB was supported by the NINDS, NIH (R21NS108138). African American Health was supported by a grant from the National Institute on Aging (R01 AG010436 to DKM). AAB and ADN also acknowledge financial support provided by Saint Louis University.

## Author contributions

AAB was responsible for data management, data analysis, conceptual development, interpreting data, and preparation of the first draft of the manuscript. GA was responsible for generating data and preliminary data analysis. ADN provided resources and supervised data acquisition. TM was responsible for managing the complete AAH dataset, provided data, and assisted with analysis. DKM and JEM were responsible for conceptual development, design, and funding acquisition for the AAH study. All authors read and approved the manuscript for content. All authors read and approved the manuscript for content.

## Competing interest’s statement

The authors have no competing interests to report.

## Notes

**Disclosures:** The authors declared no conflict of interest

### Competing Interest Statement

The authors have declared no competing interest.

### Funding Statement

AAB was supported by the NINDS, NIH (R21NS108138). AAB and ADN acknowledge financial support provided by Saint Louis University. African American Health was supported by a grant from the National Institute on Aging (R01 AG010436 to DKM).

### Author Declarations

All study procedures were approved by the Institutional Review Board at Saint Louis University and have therefore been performed in accordance with the ethical standards laid down in the 1964 Declaration of Helsinki and its later amendments.

### Summary of Updates

Manuscript revised due to publication of adropin measurements in the MAPT study. Manuscript was simplified to focus on serum adropin levels in subjects with impaired cognition. Data comparing adropin tertiles and different fructosamine levels were removed. Additional data from snRNAseq was added.

