## supplemental table 1 for "Low circulating adropin levels in late-middle aged African Americans with poor cognitive performance"

**Table S1.** Age, sex, body mass, and serum adropin concentrations of the 352 study participants.

| **Characteristics** | **Sample size (n=352)** | Mean ± Std Deviation, range |
|---|---|---|
| Age (years) | 352 | 56.6 ± 4.4, 45 to 65 |
| Females (%) | 352 | 66.7% |
| Body mass index ($kg/m^2$) | 351 | 31.3 ± 6.9, 14.5 to 55.7 |
| Serum adropin (ng/ml) | 352 | 3.24 ± 1.47, 0.15 to 8.5 |
